# Circulating Levels of Angiotensinogen, Sex, and Hormone Therapy - The Multi-Ethnic Study of Atherosclerosis (MESA)

**DOI:** 10.1101/2024.03.22.24304764

**Authors:** Karita C. F. Lidani, Patrick J. Trainor, Robert Buscaglia, Kristoff Foster, Sophia Jaramillo, Kirolos Michael, Alexander Pete Landry, Erin D. Michos, Pamela Ouyang, Erin S. Morgan, Sotirios Tsimikas, Andrew P. DeFilippis

## Abstract

**Background:** Angiotensinogen, the unique precursor of all angiotensin hormones of the Renin-Angiotensin-Aldosterone System (RAAS), is now a potential target in a novel pharmacological approach to hypertension. Investigating the factors that influence angiotensinogen levels, including sex hormones, may have important therapeutic implications.

**Methods:** Plasma angiotensinogen and sex hormones levels were measured in 5,171 Multi-Ethnic Study of Atherosclerosis (MESA) participants. Linear models were employed to determine the associations of angiotensinogen with sex hormones, and mediation analysis was performed to evaluate the effect of HT on blood pressure (BP) and hypertension through angiotensinogen.

**Results:** Angiotensinogen levels were significantly higher in postmenopausal women receiving HT (n=760) compared to women not receiving HRT (n=1,675) and in men (n=2,736). A positive association was present between angiotensinogen and estrogen levels that differed in magnitude between sexes and by HT status among postmenopausal women (women on HT: r=0.44, p< 0.0001; women not on HT: r=0.09, p=0.0002; and men: r= 0.07, p=0.0003). The type of HT formulation (estrogen or estrogen/progesterone) and its duration of use did not significantly affect angiotensinogen levels. HT indirectly increased systolic BP (β=1.24) and the odds of hypertension (OR=1.065) through its effect of increasing angiotensinogen.

**Conclusions:** A positive association was present between angiotensinogen and estrogen levels that differed by HT status. HT impacts systolic BP and hypertension indirectly by increasing angiotensinogen. This study underscores the role of angiotensinogen in hypertension, and the complex relationship between HT and hypertension.

## INTRODUCTION

Angiotensinogen is the liver-derived protein precursor of all angiotensin (Ang) peptides of the renin-angiotensin-aldosterone system (RAAS) including Ang I and Ang II ^1–3^. The RAAS is critical to the pathology of hypertension. Therapeutics that target the downstream metabolism and reception of metabolites of angiotensinogen are proven to improve clinical outcomes for patients with hypertension, and to reduce the risk of heart failure and myocardial infarction ^4–6^. However, therapies targeting the metabolism or reception of Ang peptides derived from angiotensinogen often lead to a compensatory pathway that limits therapeutic efficacy as monotherapy ^2^. This necessitates the use of a combination with other classes of drugs to achieve adequate blood pressure control in many patients.

Angiotensinogen is produced primarily by the liver, but the other components of the RAAS pathway are mainly active in the kidney. Angiotensinogen is now a target of therapy in phase 1 and 2 trials. It is proposed that targeting angiotensinogen may optimally inhibit the RAAS pathway and provide more effective and possibly safer alternative by inhibiting angiotensinogen in the liver instead of the kidney ^7,8^.

Although angiotensinogen plays an essential role in regulating RAAS, little is known about key determinants of angiotensinogen plasma levels. Estrogen may be a significant contributor to expression and activity of RAAS components through several pathways ^9–11^. Previous studies have demonstrated that circulating levels of angiotensinogen are related to sex ^12,13^, and that the administration of exogenous estrogen to premenopausal women in contraceptive formulations ^14^ and to postmenopausal women as HT ^15–17^ increases plasma concentrations of circulating angiotensinogen. However, most of these studies are limited by small sample size and were conducted in exclusively normotensive women.

The effect of natural and supplemental estrogen on angiotensinogen levels and the factors underlying how sex and sex hormones impact angiotensinogen levels remains poorly understood. The current study aimed to better delineate the complex interactions between sex, estrogen, and HT with circulating plasma levels of angiotensinogen and their effect on blood pressure/ hypertension in a community-dwelling multi-ethnic cohort.

## METHODS

### Study participants and design

The Multiethnic Study of Atherosclerosis (MESA) enrolled 6,814 participants (3,601 women and 3,213 men) with no-known clinical atherosclerotic cardiovascular disease of four ethnic backgrounds (White, Chinese, Black, and Hispanic), with age between 45-84 years from six communities in the United States (Forsyth County, NC; Northern Manhattan and the Bronx, NY; Baltimore, MD; St. Paul, MN; Chicago, IL; and Los Angeles County, CA). The design of the study has been described in detail in an earlier publication ^18^. An ancillary study was conducted by our group that measured angiotensinogen levels in MESA participants who were not included in the MESA-1000. The MESA-1000 is a random sample of approximately 1,000 participants that has undergone additional tests using baseline blood samples. To minimize depletion of these specimens, ancillary studies exclude the MESA-1000 participants, unless otherwise justified. Therefore, this study included 5,786 participants. Medical history, laboratory data, and anthropometric measurements were ascertained as described previously ^18,19^. The study was approved by the institutional review boards of the participating institutions, and all participants gave written informed consent.

Our study population (**Figure 1**) consisted of all men and postmenopausal women who had plasma angiotensinogen levels and sex hormones measured at baseline (n=5,292). Measured sex hormones included total testosterone, dehydroepiandrosterone (DHEA), and estradiol (E2). Additionally, sex hormone binding globulin (SHBG) was measured given its role in transporting hormones in circulation. We sequentially excluded 97 pre-menopausal women, and 24 women with missing information on HT use. Our analytical sample was composed of 2,435 postmenopausal women and 2,736 men.

**Figure 1:**
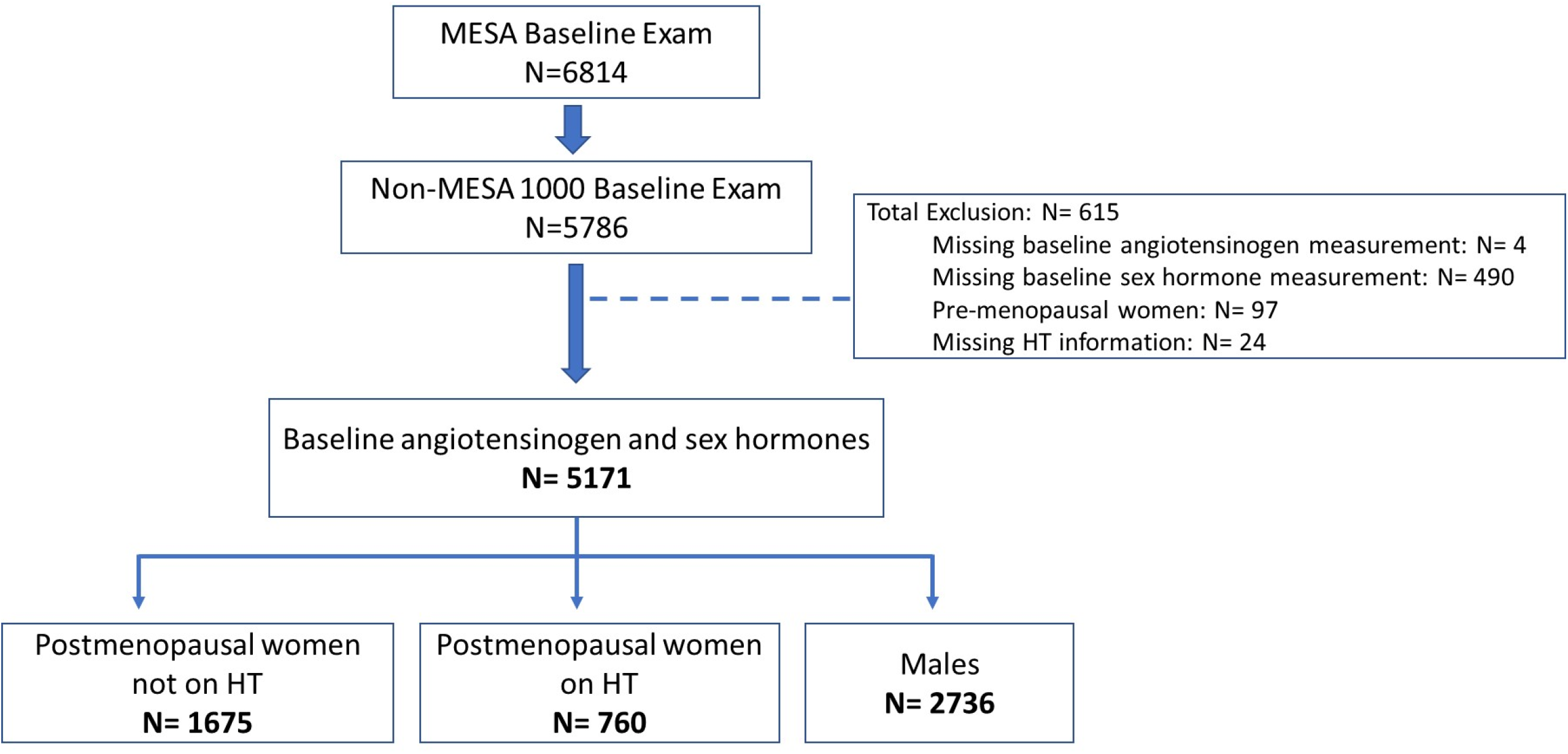
Participant inclusion/exclusion in study. HT, hormone therapy.

Menopausal status was determined through an algorithm developed using answers to a series of self-reported questions that included age, age at menopause, history of surgical menopause and age at surgical menopause as compared to age (**Supplemental Figure 1**) ^20^. Indeterminate menopausal women were not included in this analysis. Women were considered postmenopausal if they were older than 55 years of age or had undergone a bilateral oophorectomy and/or self-reported being postmenopausal or having an absence of menstrual periods in the preceding year^20^. HT consisted of estrogen alone (Premarin or Estratab) (n = 251, 33.0%) or estrogen with progesterone (Premarin plus Provera, Estratab plus Provera, Prempro, or Premphase) (n = 98, 12.9%), while 54.1% (n = 411) had no data on the type of HT. Time duration of HT use ranged from 0 to 49 years (Median = 10.0 years).

### Laboratory measurements

Circulating angiotensinogen levels and sex hormones were measured at the baseline (Exam 1) visit of the included MESA participants. Angiotensinogen measurements were made using an enzyme-linked immunoassay that has been described previously ^7^ and was executed by Medpace Reference Labs (Cincinnati, Ohio). Briefly, the coating antibody was rabbit anti-human immunoglobulin monoclonal antibody (IBL-America, Minneapolis, Minnesota). Total angiotensinogen (including intact angiotensinogen and des[Ang I] angiotensinogen) was then detected from 1:10,000 diluted EDTA plasma with horseradish peroxidase mouse anti-human angiotensinogen monoclonal Fab’ fragment (IBL-America). A standard curve was generated for quantitation using purified human angiotensinogen. The coefficient of variation was observed to be 9% with an analytical range of 13.9 µg/mL to 75.9 µg/mL.

Endogenous sex hormone levels were measured from fasting serum samples at the University of Massachusetts Medical Center in Worcester, MA. The assays and kits used to measure the hormone levels were an ultrasensitive radioimmunoassay kit for E2 (Diagnostic System Laboratories, Webster, TX), radioimmunoassay kits for total testosterone and DHEA and a chemiluminescence enzyme immunometric assay using Immulite kits for SHBG (Diagnostic Products Corporation, Los Angeles, CA) ^20–23^. The intra-assay coefficients of variation for total testosterone, SHBG, DHEA, and E2 were 12.3%, 9.0%, 11.2%, and 10.5%, respectively.

### Other covariates

Covariates were obtained from standardized questionnaires, physical exam, and laboratory measures at study visit 1 as previously reported ^18,19^. Age, race/ethnicity, smoking status, alcohol history, and age at menopause were self-reported. A medication inventory determined medication use including use of HT and antihypertensive medications at visit 1 (baseline). Diabetes was assessed by self-reported physician diagnosis, a fasting glucose level of ≥126 mg/dL, or hypoglycemic medication use. Height and weight, measured using standardized procedures, were used to calculate body mass index (BMI). Prevalent hypertension was defined as systolic blood pressure (BP) >130 mm Hg, diastolic BP >80 mm Hg, or hypertension medication use at examination 1 (baseline).

### Statistical analysis

The distribution of cohort characteristics was determined within each sex, including the use of HT in postmenopausal women. Median and quartile values (25^th^ and 75^th^ percentiles) of circulating angiotensinogen levels were determined by sex, race/ethnicity, and HT in postmenopausal women.

To determine the relationship between circulating angiotensinogen levels and sex by HT, an un-adjusted linear model was estimated by regressing levels of angiotensinogen on sex by HT and estradiol levels. This model was then refitted to incorporate other covariates that had univariable R^2^ values greater than 0.025 [race/ethnicity, BMI, total cholesterol, high-sensitivity C-reactive protein (hs-CRP), total testosterone, DHEA, and SHBG). Un-adjusted and adjusted linear models set men as the reference group, and each analyte was log-transformed and then scaled to have mean zero and standard deviation 1. The model coefficients thus reflect a one standard deviation change.

To investigate the potential mediating role of angiotensinogen in the relationship between HT use and blood pressure/ hypertension, a mediation analysis was conducted to delineate: (1) the direct effect of HT on blood pressure/ hypertension unrelated to angiotensinogen; and (2) the indirect effect of HT on blood pressure/hypertension mediated by angiotensinogen. Regression models were adjusted for age, BMI, race/ethnicity, and smoking history, while the use of any hypertension medication was included as a covariate only for the analysis of blood pressure.

The analyses reported in the current work were conducted using the R statistical language ^24^ (version 4.0.2) and the following packages: *ppcor* ^25^, *boot* ^26^, *emmeans* ^27^, *dplyr* ^28^, *ggplot2* ^29^, and *mediation* ^30^.

## RESULTS

Baseline demographics of the 5,171 participants are reported in **Table 1**, stratified by sex and the use of HT in postmenopausal women. Across all race/ethnicity groups, median angiotensinogen levels were highest in postmenopausal women on HT, followed by women participants not on therapy, followed by men (Medians: 36.0 µg/mL, 20.7 µg/mL, and 18.3 µg/mL, respectively) (**Figure 2**). The most pronounced relative difference across race / ethnicity in angiotensinogen levels was observed in the ratio between White and Chinese individuals. Postmenopausal White women on HT have an angiotensinogen ratio of 1.26 (95% CI of 1.15-1.38); followed by postmenopausal white women not on HT (ratio = 1.10 and 95% CI of 1.05-1.16); and men (ratio = 1.06 and 95% CI of 1.02-1.10) as compared to Chinese women and men **(Figure 2, Supplemental Table 1**).

**Figure 2:**
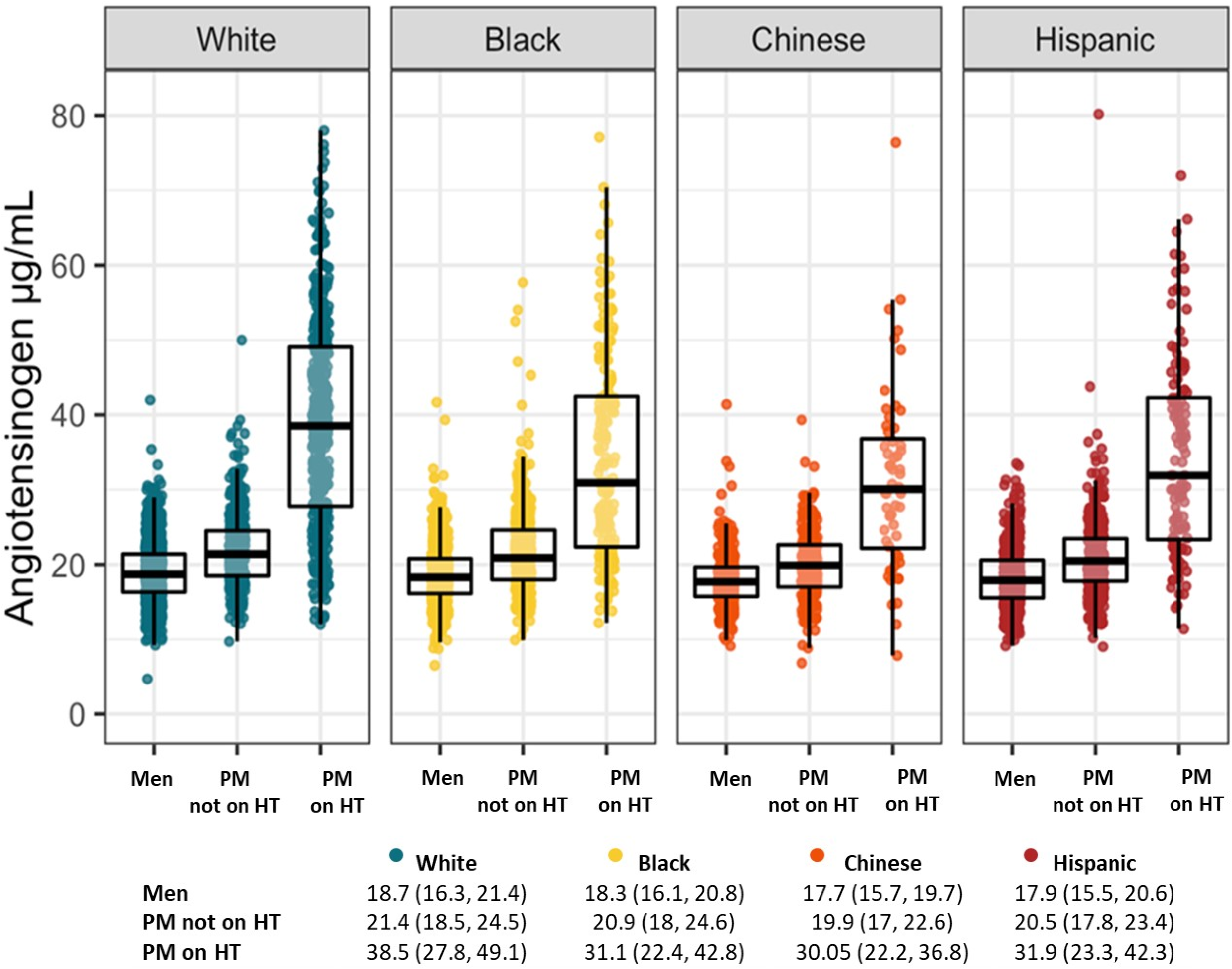
Distribution of circulating angiotensinogen levels by sex, race/ethnicity, and hormone therapy (HT) status in postmenopausal (PM) women. Boxplot shows 25^th^ percentile, median, and 75^th^ percentile. Table shows median and quartile values (25^th^ and 75^th^ percentiles).

**Table 1:** Cohort characteristics of the 5,171 MESA participants stratified by sex and Hormone Therapy (HT). Percentages presented are the number of participants in each row variable over the total number in each column.

| Characteristic | Men<br>(N = 2736) | PM not on HT<br>(N = 1675) | PM on HT<br>(N = 760) |
| --- | --- | --- | --- |
| Mean age ± SD, years | 62.6 ± 10.2 | 66.2 ± 9.2 | 62.6 ± 8.7 |
| Race/Ethnicity, n (%) |  |  |  |
| White | 1037 (37.9) | 487 (29.1) | 410 (53.9) |
| Black | 737 (26.9) | 524 (31.3) | 177 (23.3) |
| Chinese | 351 (12.8) | 248 (14.8) | 56 (7.4) |
| Hispanic | 611 (22.3) | 416 (24.8) | 117 (15.4) |
| Diabetes, n (%) | 383 (14.0) | 234 (14.0) | 65 (8.6) |
| Mean BMI ± SD, kg/m<sup>2</sup> | 27.8 ± 4.4 | 28.9 ± 6.0 | 28.0 ± 5.9 |
| Mean total cholesterol ± SD, mg/dL | 188.0 ± 35.1 | 202.7 ± 36.5 | 199.4 ± 33.5 |
| Mean HDL cholesterol ± SD, mg/dL | 45.1 ± 11.8 | 54.7 ± 14.5 | 61.2 ± 16.8 |
| Mean LDL cholesterol ± SD, mg/dL | 116.5 ± 31.1 | 122.6 ± 32.7 | 110.3 ± 29.6 |
| Mean systolic BP ± SD, mmHg | 126.1 ± 19.4 | 131.3 ± 24.0 | 126.9 ± 22.0 |
| Mean diastolic BP ± SD, mmHg | 75.1 ± 9.4 | 69.6 ± 10.4 | 68.3 ± 9.9 |
| Hypertension, n (%) | 1185 (43.3) | 873 (52.1) | 374 (49.2) |
| Any hypertension medication, n (%) | 997 (36.4) | 694 (41.4) | 327 (43.1) |
| Any lipid lowering medication, n (%) | 454 (16.6) | 325 (19.4) | 143 (18.8) |
| Current aspirin use, n (%) | 787 (28.8) | 391 (23.4) | 199 (26.2) |
| Smoking history, n (%) |  |  |  |
| Never | 1115 (40.9) | 1066 (63.8) | 365 (48.3) |
| Former | 1229 (45.1) | 432 (25.9) | 305 (40.3) |
| Current | 383 (14.0) | 172 (10.3) | 86 (11.4) |
| Alcohol history, n (%) |  |  |  |
| Never | 279 (10.3) | 601 (36.1) | 168 (22.4) |
| Former | 748 (27.5) | 360 (21.6) | 156 (20.8) |
| Current | 1690 (62.2) | 704 (42.3) | 426 (56.8) |
| Median hs-CRP [Q1, Q3], mg/L | 1.390 [0.690, 3.060] | 2.210 [0.978, 4.695] | 3.515 [1.490, 6.945] |
| Median Creatinine [Q1, Q3], mg/dL | 1.02 [0.92, 1.22] | 0.82 [0.72, 0.92] | 0.82 [0.72, 0.92] |
| Mean eGFR ± SD | 75.7 ± 16.0 | 71.8 ± 15.8 | 71.6 ± 15.0 |
| Total testosterone [Q1, Q3], nmol/L | 14.12 [11.31, 17.67] | 0.94 [0.62, 1.39] | 0.76 [0.45, 1.18] |
| Dehydroepiandrosterone [Q1, Q3], nmol/L | 12.4 [9.1, 17.0] | 10.9 [7.5, 15.2] | 8.7 [6.0, 12.7] |
| Sex Hormone Binding Globulin [Q1, Q3], nmol/L | 41.2 [31.5, 53.0] | 50.6 [37.2, 70.3] | 103.0 [63.1, 163.2] |
| Estradiol [Q1, Q3], nmol/L | 0.114 [0.088, 0.139] | 0.059 [0.040, 0.081] | 0.231 [0.128, 0.356] |

Note: data are shown as mean and standard deviation (SD), n (%), or median [Q1, Q3].
BMI, body mass index; BP, blood pressure; HDL, high-density-lipoprotein; hs-CRP, high-sensitivity C-reactive protein; LDL, low-density-lipoprotein; eGFR; Q1, first quartile (25<sup>th</sup> percentile); Q3, third quartile (75<sup>th</sup> percentile).

Positive associations between estradiol and angiotensinogen levels were observed in all participants and were of greater magnitude in postmenopausal women on HT than those not on HT and men (postmenopausal women on HT: *r* = 0.44, p < 0.0001; postmenopausal women not on HT: *r* = 0.09, p = 0.0002; and men: *r* = 0.07, p = 0.0003). In the regression model presented in **Figure 3**, the association between estradiol and angiotensinogen differs by specific group (men, postmenopausal women not on HT, and postmenopausal women on HT). Using the model, we compared angiotensinogen levels at the average estradiol levels for each group. At the average log-estradiol level for men, angiotensinogen was 18.2 μg/mL with a 95% confidence interval of (18.0, 18.4). At the average log-estradiol level for women not on HT it was 20.8 μg/mL with a CI of (20.6, 21.1). At the average log-estradiol levels for postmenopausal women on HT it was 33.9 μg/mL with a CI of (33.4, 34.5). For the same estradiol level, 0.2 nmol/L (mean for entire cohort), men had a mean circulating angiotensinogen level of 18.7 μg/mL, postmenopausal women not on HT had a mean angiotensinogen level of 19.5 μg/mL, and postmenopausal women on HT had a mean angiotensinogen level of 33.9 μg/mL (**Figure 3**).

**Figure 3:**
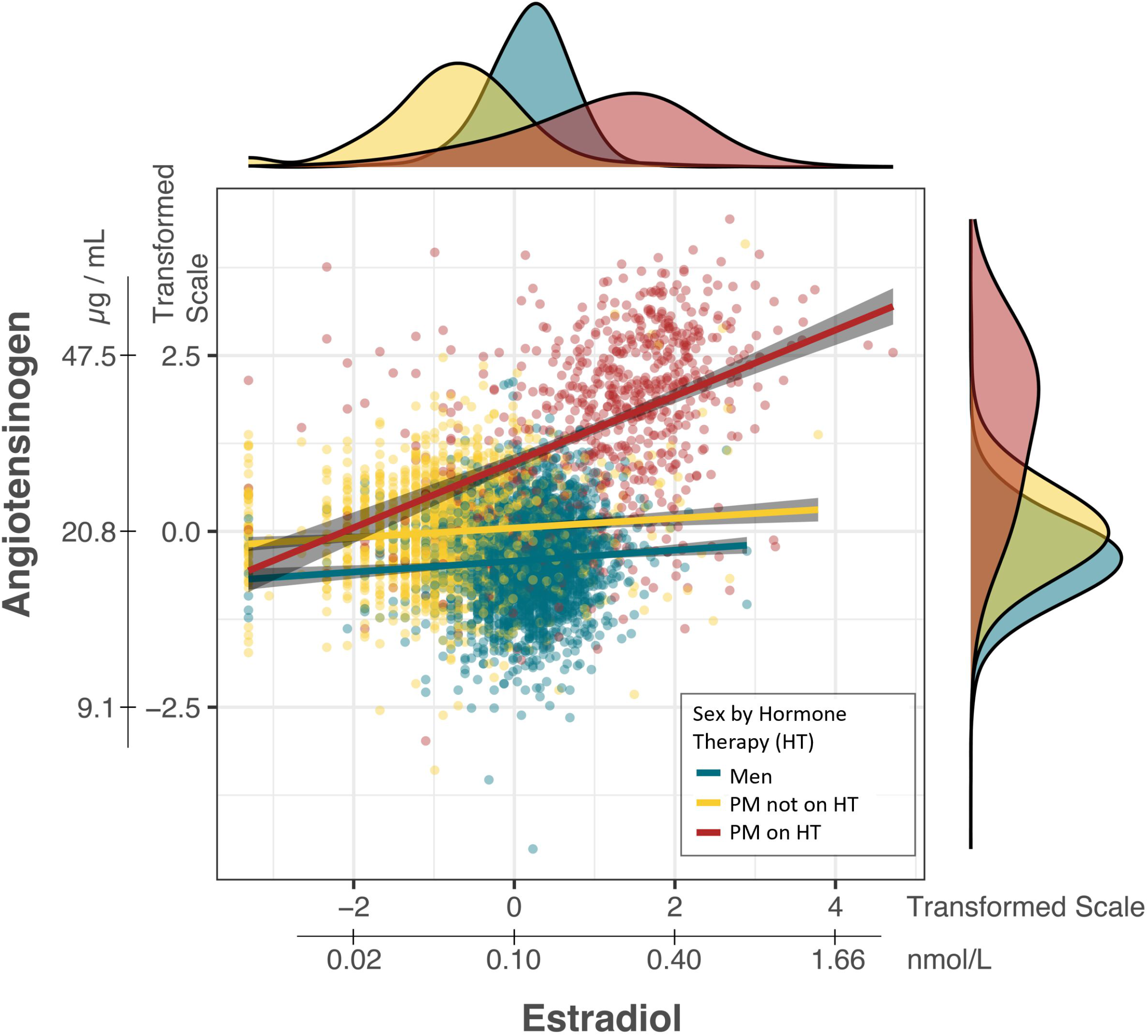
Linear relationship between log-transformed estradiol and log-transformed angiotensinogen levels stratified by sex and hormone therapy (HT) status with density curves in the margin showing the distribution of each by group. Regression lines are shown based on an interaction effect between sex and HT. Each dot represents a participant. PM, postmenopausal women.

An estimate one standard deviation (SD) higher log-estradiol level was associated with 0.195 SD, 0.585 SD, and 1.716 SD higher log-angiotensinogen (95% CI: 0.169, 0.220) in men, postmenopausal women not on HT, and postmenopausal women on HT, respectively (**Table 2**). Indicating smaller differences in angiotensinogen for the same amount of change in estradiol in men and postmenopausal women not on HT as compared to those on HT (**Table 2**, **Figure 3)**.

**Table 2:** Un-adjusted model for predicting angiotensinogen from estradiol, in men, post-menopausal women not on HT, and post-menopausal women on HT. Note: Each analyte was log-transformed and then scaled to have mean zero and standard deviation 1. The model coefficients are thus in terms of standard deviation units. Men were considered the reference level for sex.

| Variable | Estimate | (95% CI) | p-value |
| --- | --- | --- | --- |
| Intercept | -0.443 | (-0.471, -0.414) | <0.0001 |
| PM not on HT | 0.585 | (0.534, 0.636) | <0.0001 |
| PM on HT | 1.716 | (1.652, 1.781) | <0.0001 |
| Estradiol | 0.195 | (0.169, 0.220) | <0.0001 |

$R^2 = 0.435$
<sup>†</sup>Estradiol concentrations were log-transformed, centered, and standard deviation scaled.
PM: postmenopausal women.
HT: hormone therapy

To further examine if the differences in the association between estrogen and angiotensinogen between men, postmenopausal women not on HT and postmenopausal women on HT, we adjusted the model described in **Table 2** for variables with univariable model R^2^ values greater than 0.025 (**Supplemental Table 2**). Following adjustment for race / ethnicity, BMI, total cholesterol, hs-CRP, total testosterone, DHEA, and SHBG, the model coefficients for sex, HT, and estradiol changed little in terms of direction or magnitude (**Supplemental Table 3**). The proportion of the variance in angiotensinogen explained by estrogen, sex and HT status alone is 43.5%; increases to 48.0% with the addition of the variables noted above (**Table 2 and Supplemental Table 3**) and the partial *R*^2^ values for estradiol changed little following model adjustment (**Supplemental Table 4**).

Different formulations of HT, only with estrogen or combined estrogen/progesterone, showed similar levels of angiotensinogen with median value of 36.0 µg/mL (95% CI: 32.7, 40.5) and 36.7 µg/mL (95% CI: 33.7, 40.5), respectively. In addition, the duration of HT use was not associated with angiotensinogen levels.

In the mediation analysis, we examined the role of angiotensinogen in explaining the relationship between HT and blood pressure/ prevalent hypertension. The indirect effect of HT mediated through angiotensinogen levels was associated with increased systolic BP (β = 1.24, p = 0.040) and odds of hypertension (OR = 1.065, p < 0.001). The total effect of HT slightly increased the odds of hypertension (OR = 1.082, p < 0.001) (**Table 3**).

**Table 3.** Direct, indirect, and total effects of HT on systolic blood pressure (BP), diastolic BP, and hypertension with angiotensinogen as a mediator. Note: Angiotensinogen levels have been log-transformed and standardized. Models to regress angiotensinogen on HT were adjusted for age, race/ethnicity, BMI, smoking history, and use of any hypertension medication. Models to regress blood pressure/ hypertension on HT were adjusted for angiotensinogen levels, age, race/ethnicity, BMI, smoking history, and use of any hypertension medication (only for blood pressure).

| Dependent Variable | Effects | Estimate | (95% CI) | p-value |
| --- | --- | --- | --- | --- |
| Systolic BP | **Direct Effect**<br>*HT → Systolic BP* | -1.19 | (-3.36, 1.14) | 0.306 |
|  | **Indirect Effect**<br>*HT → Angiotensinogen → Systolic BP* | 1.24 | (0.357, 2.48) | 0.040 |
|  | **Total Effect** | 0.048 | (-1.88, 1.87) | 0.953 |
| Diastolic BP | **Direct Effect**<br>*HT → Diastolic BP* | -1.710 | (-2.78, -0.55) | < 0.0001 |
|  | **Indirect Effect**<br>*HT → Angiotensinogen → Diastolic BP* | 0.472 | (-0.16, 1.08) | 0.126 |
|  | **Total Effect** | -1.234 | (-2.13, -0.27) | 0.014 |
| Hypertension | **Direct Effect**<br>*HT → Hypertension* | 1.016 | (0.97, 1.06) | 0.530 |
|  | **Indirect Effect**<br>*HT → Angiotensinogen → Hypertension* | 1.065 | (1.04, 1.09) | < 0.001 |
|  | **Total Effect** | 1.082 | (1.04, 1.13) | < 0.001 |

BMI, body mass index; HT, hormone therapy; BP, blood pressure.

## DISCUSSION

The key findings of this study are as follows: (1) across all race/ethnicity groups, median angiotensinogen levels were highest in postmenopausal women on HT, as compared to postmenopausal women not on HT and men; (2) estrogen, sex and HT status alone accounts for 43.5% of variance in circulating angiotensinogen levels; (3) the indirect effect of HT mediated through angiotensinogen levels increases systolic blood pressure and the odds of hypertension, but did not have an impact on diastolic BP. These findings highlight the impact of postmenopausal HT on circulating angiotensinogen levels, and the potential effect of HT on prevalent hypertension through angiotensinogen levels.

Despite the availability of effective antihypertensive treatments, nearly half of patients with hypertension fail to achieve BP goals recommended by guidelines. The pursuit of novel antihypertensive therapies includes the development of antisense oligonucleotides (ASOs) and small interfering RNA (siRNA), designed to reduce angiotensinogen secretion ^2,3,8^. Clinical trials with ASO in Phase 1 (NCT03101878) on healthy volunteers and Phase 2 on patients with mild (NCT03714776) and uncontrolled (NCT04083222) hypertension demonstrated favorable safety and tolerability. These trials also showed targeted reductions in angiotensinogen levels, resulting in significant decreases in both systolic and diastolic BP without compromising renal function or increasing cardiovascular risk^7^. Similarly, Phase 1 trials with siRNA (NCT03934307) in uncontrolled hypertension have shown promise in reducing blood pressure without worsening renal function^8,31,32^. Phase II trials assessing efficacy and optimal dosing are currently underway, with a predicted completion by 2025. Understanding the determinants of angiotensinogen levels, including sex hormones, race, sex, and menopausal status may influence the efficacy of this new class of therapeutics.

Consistent with our results, previous studies demonstrated that postmenopausal estrogen therapy was associated with increased angiotensinogen plasma concentrations ^16,33^. The role of exogenous estrogen in stimulating *AGT* gene expression or secretion have been demonstrated in animal models and hepatic cell line ^34–36^. In addition Schunkert et al. ^16^ found increased angiotensinogen levels to a similar extent in women taking formulations that contained only estrogen or estrogen/progestin combination, both being significantly higher compared with women without HT. Although there are recognized sex differences in blood pressure regulation as well as cardiovascular disease susceptibility, onset, prevalence, clinical presentation, pathophysiology, treatment responses and outcomes, the effects of exogenous estrogen on angiotensinogen and its downstream metabolites, and consequently on blood pressure are limited ^16,37^. Trainor et al. (2023)^13^, using the same cohort of the present study, reported positive associations between angiotensinogen and blood pressure/prevalent hypertension.

The effect of HT on prevalent hypertension, evidenced by mediation analysis, indicates that HT indirectly increases the odds of prevalent hypertension via increased angiotensinogen, which contributes to the total effect of HT on hypertension (**Central Illustration**). In systolic BP, this indirect effect was counterbalanced by HT direct effects (independent of angiotensinogen) that decrease systolic BP. This data suggests that the blood pressure effects of HT may be clinically exploited by silencing the angiotensinogen mediated indirect blood pressure raising properties of HT with newly developed therapeutics that directly target angiotensinogen production ^2^.

Previous studies have evaluated the relationship between HT use and the development of hypertension. The Women’s Health Initiative I (WHI I, <u>NCT00000611</u>], a randomized controlled trial (RCT) assessing primary prevention in 16,608 participants, observed an increase in systolic blood pressure among postmenopausal women using combined estrogen/progestin therapy ^38^. This is consistent with other prospective studies in normotensive postmenopausal women ^39,40^. Conversely, the Kronos Early Estrogen Prevention Study (KEEPS, <u>NCT00154180</u>), a RCT with 727 participants reported a neutral effect of HT on systolic blood pressure ^41^. In postmenopausal women with preexisting hypertension, randomized prospective studies with sample size less than 100 demonstrated that the HT use was associated with both a decrease ^42,43^ and neutral ^44^ effects on blood pressure. Notably, the Heart and Estrogen/progestin Replacement Study I (HERS I, <u>NCT00319566</u>) (RCT, n=2763) found no significant association between HT and the incidence of myocardial infarction (MI) or stroke/transient ischemic attack in postmenopausal women with established coronary disease ^45^. However, the WHI I study observed higher stroke rates in the HT group compared to the placebo ^38^. A meta-analysis of 19 randomized controlled trials, encompassing data from 40,410 postmenopausal women, furnished evidence that HT does not affect the incidence of MI for either primary or secondary prevention but lead to a greater risk of strokes ^46^. Given that HT status was one of the major determinants of angiotensinogen levels in our study, further investigations are warranted to elucidate the specific interactions between HT, angiotensinogen, and the development of hypertension and hypertension-related cardiovascular diseases (e.g., stroke).

### Study limitations

There are limitations to this study. First, we restricted our sample to postmenopausal women as both sex hormone levels and cardiovascular risk differ between pre- and postmenopausal women, and there were relatively few pre-menopausal women in MESA (17%), which would limit analyses in this subset. Second, route and dosage of HT therapy was not available for this analysis, both of which may impact on angiotensinogen levels. Third, sex hormones in MESA were not measured using the current gold standard of mass spectrometry but rather using radioimmunoassay and thus might be prone to measurement error.

## CONCLUSIONS

Angiotensinogen and estrogen levels showed a positive correlation that differ in magnitude between sexes, with the strongest association evidenced in postmenopausal women on HT. We observed evidence the HT indirectly increase systolic BP and odds of hypertension by increasing angiotensinogen while lowering systolic BP via angiotensinogen independent direct effects. This furthers our understanding of the implications of HT on angiotensinogen and BP in postmenopausal women which may influence the utility of a new class of therapeutics designed to directly reduce the production of angiotensinogen.

## DATA AVAILABILITY

Interested investigators may request access to the data by contacting the MESA data coordinating center.

## Graphical Abstract

Association between angiotensinogen and sex, and its clinical implications in postmenopausal women on hormone therapy (HT).

